# Large-scale pharmacokinetic reconstruction of propofol effect-site concentrations during anaesthetic induction

**DOI:** 10.64898/2026.03.04.26347547

**Authors:** Brent Ershoff

**Author notes:** **Corresponding author:** Brent Ershoff.

## Abstract

**Background:** Propofol dosing guidelines recommend age-based reductions because hypnotic sensitivity increases in older adults. Most real-world evaluations of induction practice, however, have relied on total weight-normalized dose (mg kg^−1^) rather than estimating cerebral exposure using pharmacokinetic models. Because age-related pharmacokinetic changes alter the relationship between administered dose and peak effect-site concentration (*C_e,max_*), mg kg^−1^ surrogates may misrepresent true age-dependent exposure during induction.

**Methods:** A retrospective reconstruction of 250,640 adult anesthetic inductions was performed using high-fidelity EHR medication timestamps. Propofol effect-site conrcentration trajectories were simulated at 1-second resolution using the Eleveld model. *C_e,max_* was benchmarked against age-adjusted hypnotic requirements (*Ce*_50_) derived from the Eleveld model. Age–exposure relationships were estimated using covariate-adjusted natural cubic splines, controlling for BMI, sex, and ASA physical status.

**Results:** From young adulthood (18–24 years) to the oldest cohort (85–89 years), weight-normalized induction doses were reduced by 32% (3.16 to 2.16 mg kg^−1^). However, modeled *C_e,max_* declined by only 17% (3.70 to 3.06 *µ*g ml^−1^), while the estimated physiological requirement declined by 34% (3.37 to 2.21 *µ*g ml^−1^), creating a widening titration offset with age. At age 75, the adjusted probability of exceeding the individual hypnotic requirement was 89.6% (95% CI: 89.3–89.8%). Notably, 54.7% (95% CI: 54.2–55.2%) of 75-year-old patients achieved peak exposures exceeding the average requirement of a healthy 20-year-old, indicating persistent anchoring of exposure to youthful levels. Findings were robust across model specifications and inclusion criteria.

**Conclusions:** In over a quarter-million inductions, real-world age-based dose reductions did not produce proportional reductions in peak propofol brain exposure. Achieved concentrations declined far more slowly than modeled geriatric sensitivity increases, consistent with systematic over-exposure in older adults. These findings suggest that weight-based dosing heuristics inadequately capture age-dependent exposure and support a transition toward exposure-informed and neurophysiologically guided induction titration in geriatric anesthesia.

## INTRODUCTION

Older adults represent a rapidly expanding surgical population with millions undergoing procedures that require general anesthesia each year^1–3^. Aging is accompanied by clinically important pharmacodynamic changes in hypnotic sensitivity: for a given propofol exposure, older patients achieve loss of consciousness at significantly lower effect-site concentrations than younger cohorts, reflecting a profound age-dependent shift in neural sensitivity.^4^. Classic dose-finding studies established a monotonic decline in the propofol effect-site concentration required for loss of consciousness (*C_e_*_50_) as a function of age—a relationship now codified in contemporary pharmacokinetic-pharmacodynamic (PK-PD) frameworks used for target-controlled infusion^5^. These pharmacodynamic observations underpin widely taught recommendations to reduce propofol dosing in older adults and are reflected in the product label that recommends lower induction doses in geriatric patients, in part to mitigate hypotension, excessive hypnotic depth, and downstream complications^6^.

Despite strong physiologic rationale, induction practice in the real-world may not consistently translate age-related pharmacodynamic knowledge into bedside titration. Previous audits and registry studies suggest that many older adults receive induction doses that exceed the recommended ranges on the label^7–10^. However, a critical unresolved question remains: does bedside dosing—even when ostensibly reduced—produce an effect-site concentration trajectory that mirrors the known decline in physiologic requirement? While clinicians may down-titrate absolute doses for older adults, the magnitude of these reductions may fail to keep pace with the exponential increase in hypnotic sensitivity. In other words, a reduction in the mass of drug administered may not yield a commensurate reduction in brain exposure, potentially masking a systematic overshoot in geriatric patients.

A key limitation of many prior studies is the reliance on static exposure metrics, such as weight-based dose (mg kg^−1^), as the primary measure of propofol effect^7^. Static metrics ignore the temporal resolution of drug delivery; a single rapid bolus and a fractionated induction may involve identical cumulative doses yet produce profoundly different peak effect-site concentrations. Furthermore, simple weight-based scaling fails to account for the dynamic covariate structures within validated pharmacokinetic models. In these frameworks, age-and body-composition-dependent changes in compartment volumes and clearances fundamentally alter the trajectory of the effect-site concentration. Consequently, dose-based audits likely provide an incomplete—and potentially misleading—estimate of anesthetic exposure, that may underestimate the degree of clinically significant overexposure in the elderly.

To evaluate whether clinical practice achieves age-appropriate propofol exposure, it is necessary to evaluate concentration rather than dose. While effect-site concentration is directly available in prospective target-controlled infusion (TCI) studies^11,12^, such information is rarely available in large retrospective cohorts dominated by manual bolus dosing and heterogeneous administration techniques. In this study, high-resolution electronic medication timestamps were leveraged to reconstruct longitudinal, patient-specific propofol effect-site concentration trajectories at an unprecedented scale. The primary objective was to characterize the relationship between patient age and the maximum estimated effect-site concentration (*C_e,_*_max_) achieved during induction, comparing these observed exposures to the age-adjusted *C_e_*_50_ requirements predicted by established pharmacodynamic frameworks.^13^. The analysis additionally evaluated whether conventional mg kg^−1^ dosing trends mirror modeled changes in effect-site exposure, testing the hypothesis that age-related reductions in administered induction doses are disproportionate to—and ultimately insufficient to offset—the pharmacokinetic and pharmacodynamic shifts that dictate actual cerebral exposure in the elderly.

## METHODS

### Study design and data source

This retrospective observational study was reviewed by the UCLA Institutional Review Board (IRB# 26-0331) and determined to be exempt because it did not meet the regulatory definition of human subjects research. All analyses used a de-identified dataset derived from the institutional perioperative data warehouse, which contains anesthesia documentation, medication administration records, and physiologic timestamps from the electronic medical record^14^.

### Study population and exclusions

Adult surgical cases (age *≥* 18 years) receiving general anaesthesia with propofol were identified from all eligible cases recorded in the perioperative data warehouse from March 2013 through the most recent de-identified data refresh available at the time of extraction in February 2026. Of 258,741 initial cases, those with documented dose errors, missing simulation variables, or physiologic outliers were excluded sequentially (Figure 1). Because the source system records all patients aged *≥* 90 years as age 90, the primary analysis was restricted to patients aged 18–89 years, yielding a final analytic cohort of 250,640 cases. Analyses including the administratively masked *≥* 90-year-old cohort were performed as sensitivity analyses.

**Figure 1:**
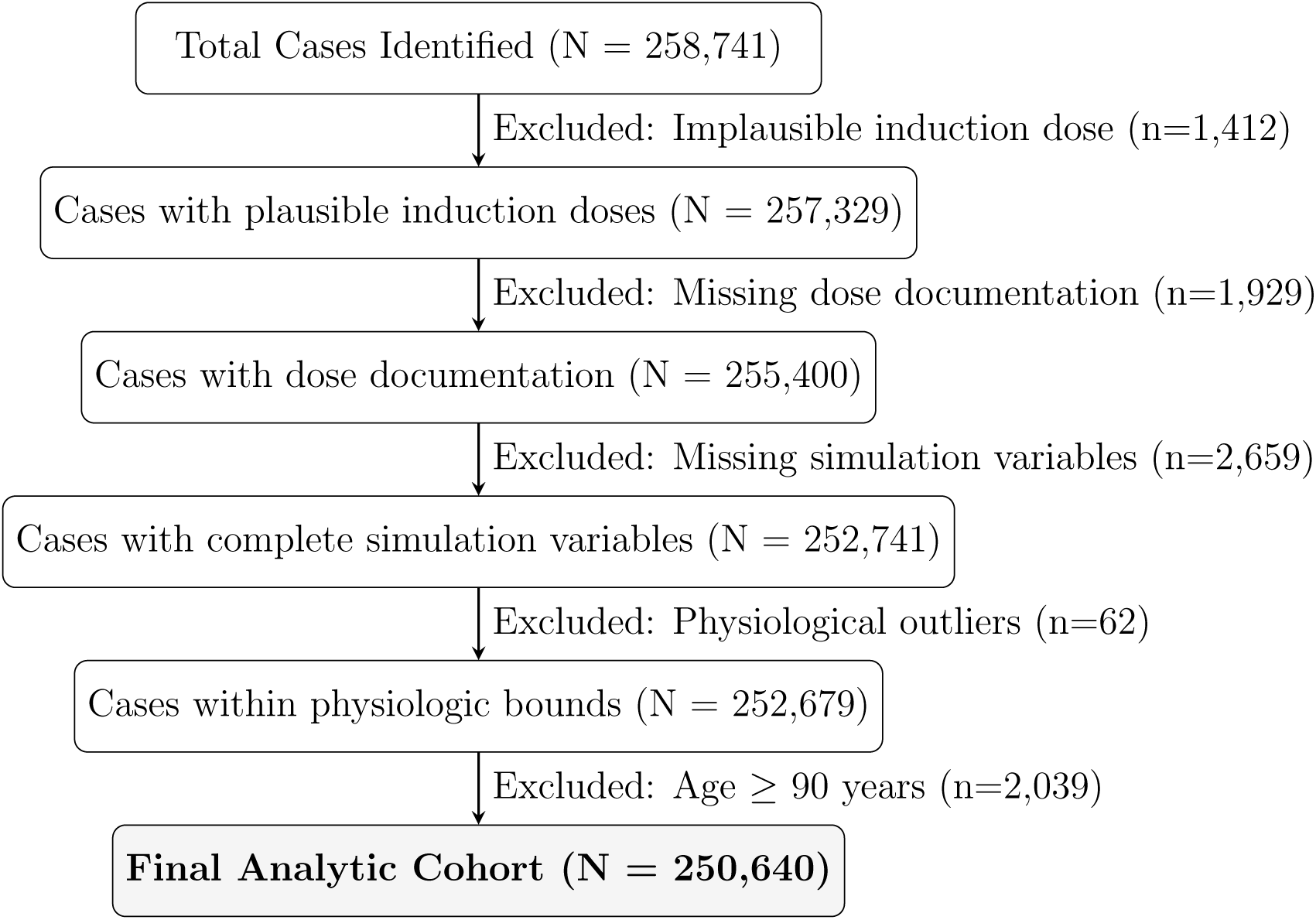
Study flow diagram. Cohort derivation for the primary analytic sample used in the pharmacokinetic reconstruction and regression analyses. Sequential exclusions were applied for implausible induction dosing, missing dose documentation, missing simulation variables, physiologic outliers, and administratively masked age *≥* 90 years.

### Data extraction and dosing metrics

Propofol administration records were retrieved using Structured Query Language (SQL) from cases in which the documented induction event preceded the intubation timestamp. For each included case, the induction timestamp served as the reference point, and all propofol administrations within 10 minutes were extracted. These included bolus doses (mg) and infusions characterized by start/stop times and rate changes (*µ*g kg^−1^ min^−1^).

Weight-normalized induction dose (mg kg^−1^) was calculated using adjusted body weight (ABW). Ideal body weight (IBW) was calculated by the Devine formula. For patients with total body weight (TBW) *>* 120% of IBW, ABW was defined as:

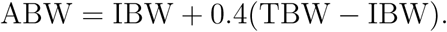

For all other patients, TBW was used as the dosing weight. Total propofol dose within the extraction window was then normalized to ABW.

### Pharmacokinetic reconstruction

Effect-site concentration time series were reconstructed by simulating validated propofol PK/PD models forward using observed medication events as input. Simulations were implemented with PyTCI^15^. The Eleveld model was used for the primary analysis because it incorporates age and body-composition covariates across a broad range of ages and body sizes^13^. Patient-specific parameters were derived from age, sex, height, and weight.

For each case, medication events were aligned to the first recorded propofol administration (*t* = 0). Concentrations were simulated at 1-second resolution for 900 seconds. Bolus doses recorded in mg were applied instantaneously. Infusion rates recorded in (*µ*g kg^−1^ min^−1^) were converted to (mg s^−1^) and applied continuously until updated by a subsequent rate change or stop event. The primary exposure metric was the peak modeled effect-site concentration during the 900-second window:

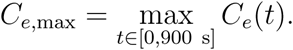

Additional implementation and reproducibility details are provided in the Supplementary Appendix.

Sensitivity analyses repeated the reconstruction using the Schnider model^16^, repeated the primary analyses without adjustment for BMI, sex, or ASA physical status, included sensitivity analyses using all documented propofol administrations, with missing documented dose values coded as 0 mg, and repeated the analysis including the administratively masked *≥* 90-year-old cohort. Additional sensitivity analyses adjusted for fentanyl, ketamine, midazolam, and etomidate administered during the induction window. Expanded sensitivity-analysis specifications are provided in the Supplementary Appendix.

### Statistical analysis and visualization

Statistical analyses were performed in Python using pandas, NumPy, statsmodels, and patsy^17–21^. Continuous outcomes, including peak effect-site concentration (*C_e,_*_max_) and weight-normalized induction dose, were modeled using ordinary least squares regression with age represented by a B-spline with five degrees of freedom. Binary outcomes, including the probability of exceeding pharmacodynamic benchmarks, were modeled using logistic regression with B-spline terms for age. Because this was a retrospective study using all eligible cases available in the perioperative data warehouse during the study period, no prospective sample-size calculation was performed.

Adjusted age trajectories were estimated using marginal predictions for a standardized patient profile in which BMI, ASA physical status, and sex were held at cohort-level means. For display, results were shown as continuous spline curves and as model-predicted values with 95% confidence intervals at the mean age of each prespecified age stratum (18–24, 25–34, 35–44, 45–54, 55–64, 65–74, 75–84, and 85–89 years).

### Pharmacodynamic reference and divergence analysis

Modeled *C_e,_*_max_ values were compared with the age-adjusted *Ce*_50_ relationship derived from the propofol-only Eleveld model:

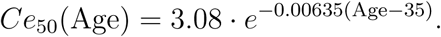

This relationship was used as a standardized physiologic reference for age-related hypnotic requirement. Divergence between modeled exposure and this reference was evaluated in three ways. First, *C_e,_*_max_, weight-normalized dose (mg kg^−1^ ABW), and *Ce*_50_ were indexed to the 18–24-year-old cohort to compare relative decline across age. Second, overexposure was defined as *C_e,_*_max_ *> Ce*_50_(Age), and the age-dependent probability of overexposure was modeled using logistic regression with B-spline terms for age, adjusted for sex, BMI, and ASA status. Third, the probability that *C_e,_*_max_ exceeded the model-predicted *Ce*_50_ for a 21-year-old (3.37 *µ*g ml^−1^) was calculated as a young-adult benchmark. For both binary outcomes, adjusted risk curves were generated by evaluating models at cohort-level means for all covariates and were displayed alongside raw observed proportions within each age stratum.

## RESULTS

A total of 258,741 anesthetic cases were identified from the UCLA perioperative data warehouse. After applying eligibility criteria, including age *≥* 18 years and anesthetic duration *≥* 30 minutes, and excluding cases with missing simulation variables or physiologically implausible anthropometrics, 250,640 unique cases comprised the final analytic cohort (Figure 1). Baseline demographic and clinical characteristics are summarized in Table 1.

**Table 1:** Baseline characteristics and induction practice by age stratum. Demographic, anthropometric, and induction-related variables for the primary analytic cohort across eight prespecified age strata. Values are presented as mean (SD) or N (%).

Part A: Ages 18–55
|  | 18-24 (N=15,004) | 25-34 (N=27,208) | 35-44 (N=33,610) | 45-54 (N=39,770) |
| --- | --- | --- | --- | --- |
| Age (Years) | 21.0 (2.0) | 29.8 (2.9) | 39.6 (2.9) | 49.7 (2.9) |
| Weight (kg) | 72.4 (20.6) | 77.5 (22.3) | 80.4 (22.8) | 81.2 (21.7) |
| Body Mass Index (kg/m<sup>2</sup>) | 25.0 (6.3) | 26.8 (7.0) | 27.8 (7.1) | 28.1 (6.7) |
| Female Sex, N (%) | 7,404 (49.3%) | 14,816 (54.5%) | 19,623 (58.4%) | 22,292 (56.1%) |
| ASA Physical Status |  |  |  |  |
| ASA 1 | 32.9% | 23.2% | 13.9% | 6.1% |
| ASA 2 | 41.1% | 46.1% | 51.0% | 48.6% |
| ASA 3 | 22.3% | 26.1% | 30.0% | 38.6% |
| ASA 4 | 3.1% | 4.0% | 4.6% | 6.0% |
| Infusion Administered, N (%) | 1,836 (12.2%) | 3,516 (12.9%) | 4,776 (14.2%) | 5,707 (14.4%) |
| >1 Bolus During Induction, N (%) | 7,267 (48.4%) | 12,659 (46.5%) | 15,291 (45.5%) | 17,650 (44.4%) |

Part B: Ages 55–89
|  | 55-64 (N=51,753) | 65-74 (N=52,886) | 75-84 (N=25,969) | 85-89 (N=4,440) |
| --- | --- | --- | --- | --- |
| Age (Years) | 59.6 (2.9) | 69.2 (2.8) | 78.6 (2.7) | 86.6 (1.4) |
| Weight (kg) | 80.6 (20.7) | 78.0 (19.0) | 74.1 (16.9) | 68.5 (15.1) |
| Body Mass Index (kg/m<sup>2</sup>) | 27.7 (6.3) | 27.1 (5.8) | 26.2 (5.1) | 24.9 (4.7) |
| Female Sex, N (%) | 25,416 (49.1%) | 24,767 (46.8%) | 12,022 (46.3%) | 2,174 (49.0%) |
| ASA Physical Status |  |  |  |  |
| ASA 1 | 2.1% | 0.6% | 0.1% | 0.0% |
| ASA 2 | 39.9% | 31.6% | 21.1% | 12.3% |
| ASA 3 | 49.1% | 58.3% | 68.7% | 74.3% |
| ASA 4 | 8.2% | 8.8% | 9.3% | 12.7% |
| Infusion Administered, N (%) | 7,840 (15.1%) | 8,323 (15.7%) | 3,689 (14.2%) | 592 (13.3%) |
| >1 Bolus During Induction, N (%) | 22,299 (43.1%) | 23,196 (43.9%) | 11,737 (45.2%) | 1,913 (43.1%) |

### Modeled effect-site concentrations (primary outcome)

To isolate the independent association of age, predicted peak effect-site concentrations (*C_e,_*_max_) and induction doses (mg kg^−1^ ABW) were generated for a standardized population profile. In covariate-adjusted analyses, modeled *C_e,_*_max_ remained above the age-adjusted *Ce*_50_ reference across all age strata (Figure 2A). Across the adult lifespan, the *Ce*_50_ benchmark declined by 34%, from 3.37 *µ*g ml^−1^ in the 18–24 cohort to 2.22 *µ*g ml^−1^ in the 85–89 cohort, whereas modeled *C_e,_*_max_ declined by 17%, from 3.70 to 3.06 *µ*g ml^−1^. Over the same age range, clinician-administered induction dose declined by 32%, from 3.16 to 2.16 mg kg^−1^ (Figure 2B).

**Figure 2:**
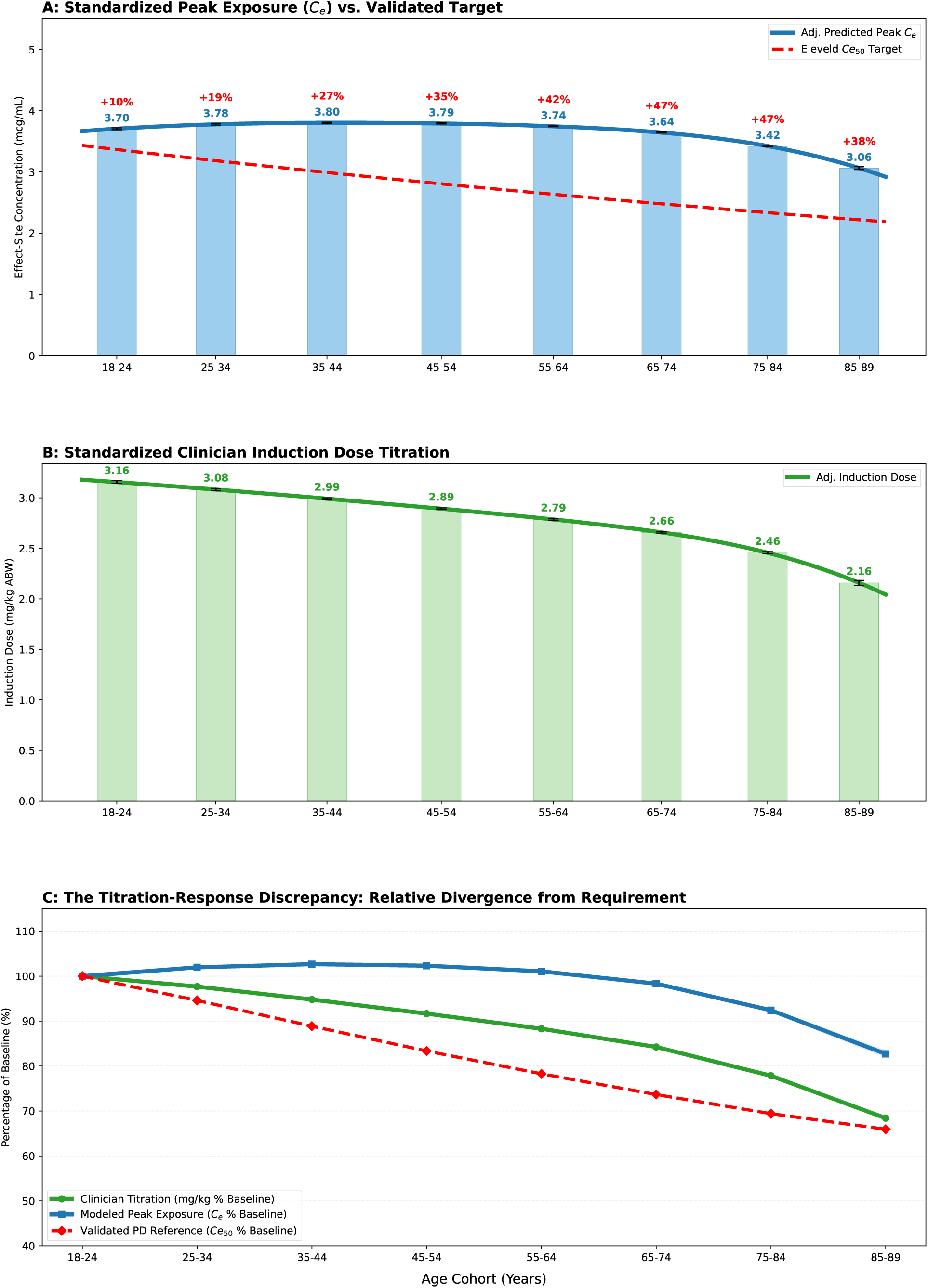
Multidimensional analysis of propofol induction dynamics. **Figure 2: Divergence of clinical dosing from modeled brain exposure and age-adjusted pharmacodynamic requirement. (A) Exposure versus requirement:** Covariate-adjusted predicted peak effect-site concentrations (*C_e,_*_max_) shown alongside age-adjusted *Ce*_50_ benchmarks. **(B) Clinical dosing trajectory:** Covariate-adjusted predicted weight-normalized induction doses (mg kg*^−^*^1^) across the adult lifespan. **(C) Normalized relative divergence:** Relative change in clinical dosing (green), modeled exposure (blue), and pharmacodynamic requirement (red), indexed to the young-adult baseline (ages 18–24). **Note:** Curves represent adjusted predicted means from spline-based regression models, with BMI, ASA physical status, and sex held at population means. Bars and error bars denote age-stratified adjusted predictions with 95% confidence intervals. In panel C, the blue confidence band denotes the 95% confidence interval for normalized modeled exposure. The dashed red line indicates the age-adjusted *Ce*_50_ benchmark from the propofol-only Eleveld model. Percentage labels indicate mean exposure surplus relative to the pharmacodynamic benchmark.

In patients aged 75–84, the adjusted mean induction dose was 2.46 mg kg^−1^ ABW, a level that remains above many commonly cited geriatric induction recommendations despite substantial age-related increases in hypnotic sensitivity. When all variables were normalized to the 18–24-year-old baseline, clinician dose declined to 68% of the young-adult value, whereas modeled *C_e,_*_max_ remained at 83% of the young-adult value (Figure 2C). Comprehensive age-stratified estimates for induction dose, modeled *C_e,_*_max_, and corresponding 95% confidence intervals are provided in Table 2.

**Table 2:** Age-stratified induction dose, modeled peak exposure, and pharmacodynamic requirement. Covariate-adjusted weight-normalized induction doses (mg kg^−1^) and modeled peak effect-site concentrations (*C_e,_*_max_, *µ*g ml^−1^) across the adult lifespan, shown alongside age-adjusted *Ce*_50_ benchmarks. Values in parentheses represent 95% confidence intervals. Induction doses are standardized to mean ASA physical status, BMI, and sex. “Surplus %” denotes the percentage by which modeled peak *C_e_*exceeds the age-adjusted *Ce*_50_ benchmark.

| Age Cohort | N | Induction Dose (mg/kg) | Peak $C_e$ (95% CI) | Target $Ce_{50}$ | Surplus (95% CI) |
| --- | --- | --- | --- | --- | --- |
| 18-24 | 14,941 | 3.16 (3.14–3.17) | 3.70 (3.69–3.72) | 3.37 | 10.1% (9.6–10.5%) |
| 25-34 | 27,088 | 3.08 (3.07–3.09) | 3.78 (3.77–3.79) | 3.18 | 18.6% (18.3–19.0%) |
| 35-44 | 33,469 | 2.99 (2.98–3.00) | 3.80 (3.79–3.81) | 2.99 | 27.2% (26.9–27.4%) |
| 45-54 | 39,583 | 2.89 (2.88–2.90) | 3.79 (3.78–3.80) | 2.80 | 35.1% (34.8–35.4%) |
| 55-64 | 51,510 | 2.79 (2.78–2.79) | 3.74 (3.74–3.75) | 2.63 | 42.1% (41.8–42.4%) |
| 65-74 | 52,594 | 2.66 (2.65–2.67) | 3.64 (3.63–3.65) | 2.48 | 46.9% (46.6–47.2%) |
| 75-84 | 25,816 | 2.46 (2.45–2.47) | 3.42 (3.41–3.43) | 2.34 | 46.6% (46.1–47.1%) |
| 85-89 | 4,419 | 2.16 (2.13–2.18) | 3.06 (3.04–3.09) | 2.22 | 38.1% (36.9–39.3%) |

### Probability of over-exposure and benchmark comparison

The probability that an individual’s modeled peak effect-site exposure (*C_e,_*_max_) exceeded their age-specific *Ce*_50_ requirement was then quantified. After adjustment for sex, BMI, and ASA physical status, logistic regression showed that the probability of over-exposure was high across older ages and generally increased across the adult lifespan before plateauing slightly at the oldest ages (Figure 3). As an illustrative geriatric example, for a standardized patient at age 75, the adjusted probability of *C_e,_*_max_ exceeding the age-specific *Ce*_50_ benchmark was 89.6% (95% CI: 89.3–89.8%). A secondary benchmark analysis evaluated the probability that an older patient’s *C_e,_*_max_ exceeded the young-adult *Ce*_50_ threshold (3.37 *µ*g ml^−1^). This probability decreased with age but remained substantial. Using age 75 as an illustrative geriatric reference point, the adjusted probability of exceeding the young-adult benchmark was 54.7% (95% CI: 54.2–55.2%).

**Figure 3:**
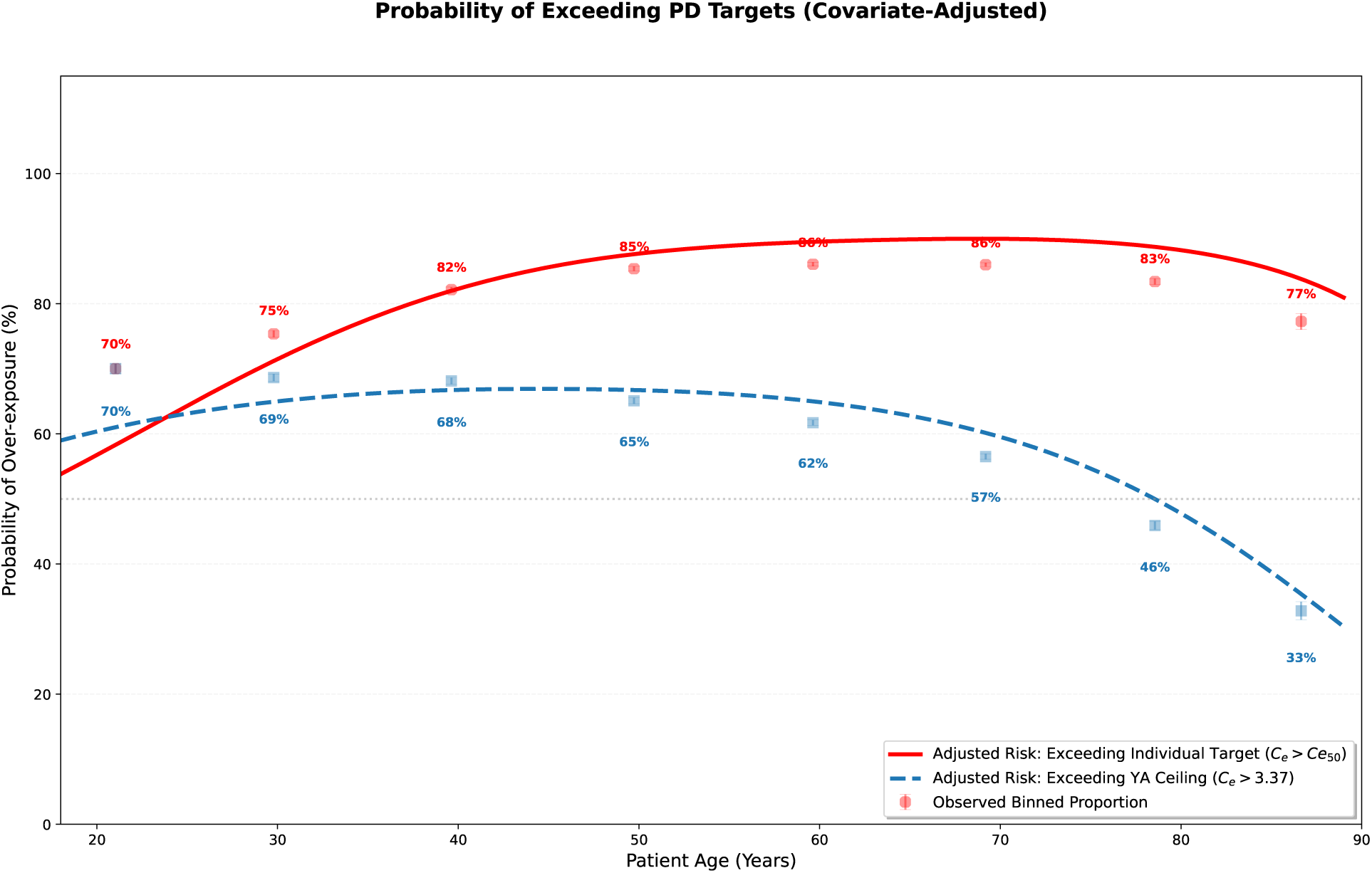
Covariate-adjusted probability of exceeding pharmacodynamic benchmarks. The red solid line shows the adjusted predicted probability that modeled peak effect-site concentration exceeds the individualized, age-specific pharmacodynamic benchmark (*C_e,_*_max_ *> Ce*_50_). The blue dashed line shows the adjusted predicted probability of exceeding the young-adult benchmark, defined as the *Ce*_50_ for the 18–24-year-old cohort. **Note:** Curves were derived from logistic regression models with age represented by spline terms and with BMI, ASA physical status, and sex held at population means. Binned points and error bars represent observed proportions with 95% Wilson score confidence intervals. Percentage labels indicate raw observed probabilities within each age stratum.

### Validation and sensitivity analyses

To assess simulation fidelity, reconstructed *C_e_* trajectories were compared with Tiva-Trainer benchmarks for representative induction sequences, including single-bolus and bolusplus-infusion scenarios^22^. Across representative benchmark scenarios, reconstructed trajectories and peak values showed close agreement with TivaTrainer outputs (Figure 4), which also summarizes the corresponding dosing schedules and peak *C_e_* values.

**Figure 4:**
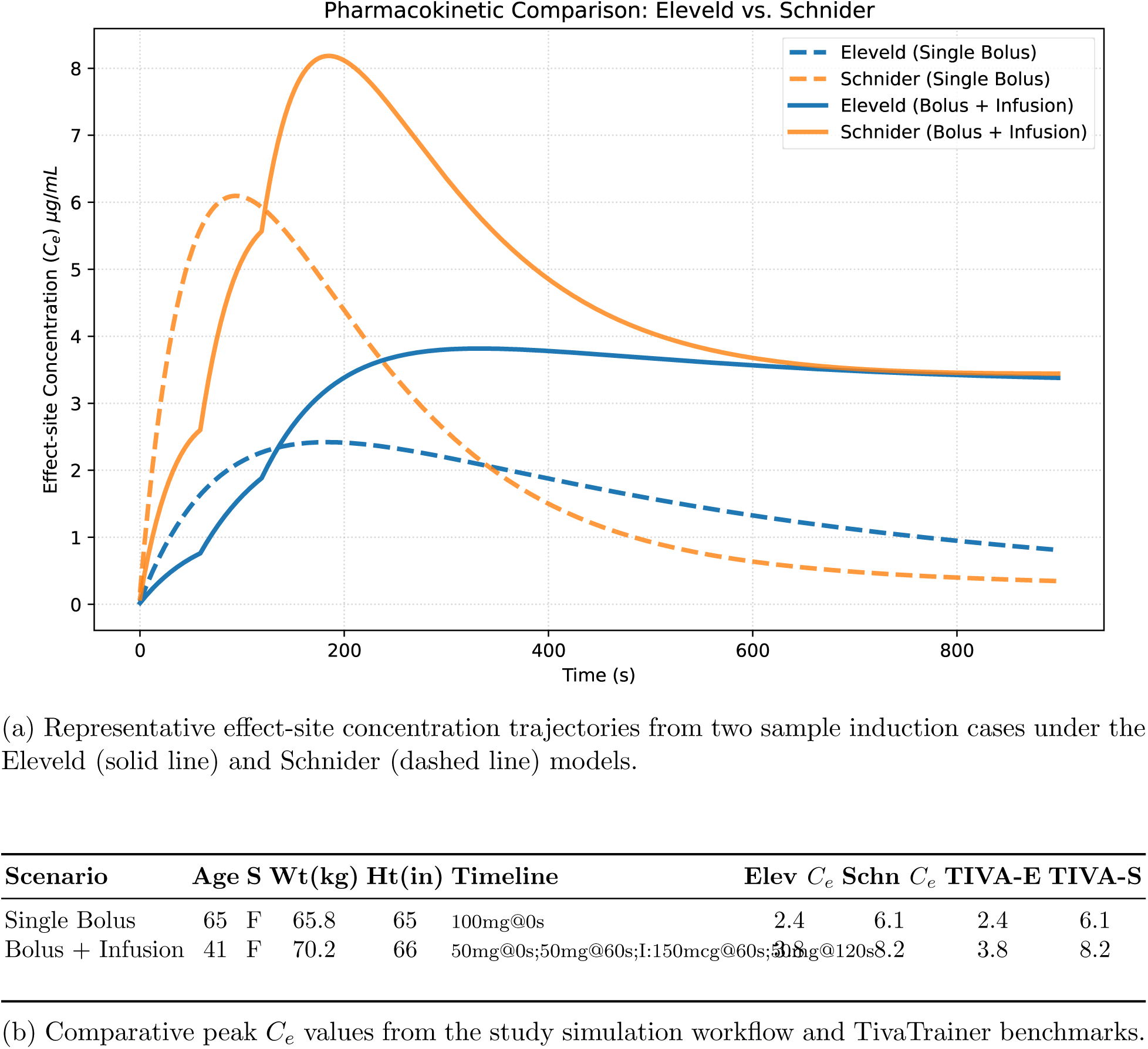
Validation of the pharmacokinetic reconstruction workflow against TivaTrainer. **(A)** Representative effect-site concentration trajectories from two sample induction cases drawn from the analytic dataset, shown under the Eleveld and Schnider models. **(B)** Comparison of peak modeled effect-site concentrations generated by the study work-flow with corresponding Tiva Trainer benchmarks across representative test scenarios. **Elev** *C_e_*and **Schn** *C_e_*denote peak concentrations calculated by the study simulation workflow under the Eleveld and Schnider models, respectively; **TIVA-E** and **TIVA-S** denote the corresponding TivaTrainer benchmarks.

### Robustness of the age-dependent divergence

The observed divergence between modeled *C_e,_*_max_ and age-adjusted pharmacodynamic requirement remained consistent across multiple sensitivity analyses:

- **PK model specification:** Repeating the primary analysis using the Schnider model^16^ yielded qualitatively similar age-associated exposure patterns, with modeled exposure declining less than clinician dose across older age groups despite age-related dose reduction (Figure S1).
- **Unadjusted analysis:** Re-estimation without covariate adjustment for BMI, sex, or ASA physical status yielded similar results (Figure S2).
- **Age-grouping sensitivity:** Inclusion of the administratively masked *≥* 90-year-old cohort did not materially alter the primary findings (Figure S3).
- **Dose inclusion thresholds:** Repeating the analysis using all documented propofol administrations, including cases initially flagged as outliers or cases with missing dose values imputed as 0 mg, yielded similar results (Figure S4).
- **Co-administered induction medications:** Additional sensitivity analyses incorporating fentanyl, ketamine, midazolam, and etomidate administered during the induction window as covariates yielded similar results (S5).

The complete age-stratified summary of sensitivity analyses, including effect estimates and 95% confidence intervals for alternative model specifications, is provided in Table S1.

## DISCUSSION

In this large-scale pharmacokinetic reconstruction of more than 250,000 anesthetic inductions, age-related reductions in propofol dose were not matched by comparable reductions in modeled peak effect-site concentration. Although clinicians administered lower doses to older adults, the resulting decline in *C_e,_*_max_ was modest relative to the marked age-related decline in hypnotic requirement. These findings suggest that routine weight-normalized dose adjustment only partially accounts for the pharmacologic changes of aging and may leave older patients exposed to induction intensities closer to those of younger adults than intended.

A central implication of this work is that the observed mismatch appears to arise from two related mechanisms. First, absolute weight-normalized induction doses in older adults often remain high relative to commonly cited geriatric dosing recommendations. Second, even when clinicians do reduce mg kg^−1^ dose with age, age-related changes in distribution volumes, inter-compartmental kinetics, and clearance attenuate the resulting reduction in brain exposure. As a result, dose reductions that appear directionally appropriate on a weight basis may still translate into substantially smaller reductions in modeled effect-site concentration than intended. Importantly, this pattern was not confined to the oldest patients. Modeled peak effect-site concentration remained relatively stable across several decades of adulthood, whereas the reference requirement declined progressively, suggesting that the divergence begins well before extreme old age. By reconstructing *C_e_*trajectories from real-world induction sequences, this study provides an exposure-based view of induction practice that is not captured by dose metrics alone.

### Comparison to prior literature and novelty

Previous studies have shown that propofol dosing in older adults often exceeds geriatric weight-based recommendations^4^. The present study extends that literature in several ways. A distinctive feature of this work is the reconstruction of modeled effect-site concentration from retrospective induction dosing data at population scale, allowing age-related differences in achieved brain exposure to be examined directly rather than inferred from administered dose. In addition, the analysis captures the dynamic complexity of real-world induction by reconstructing cumulative exposure from fragmented boluses and infusion starts rather than assuming a single static bolus. Finally, it characterizes the divergence between delivered exposure and modeled requirement across the adult lifespan rather than across broad age categories alone. Together, these features provide a more detailed description of how clinical titration departs from age-related physiology.

### Clinical implications

These findings have potential implications for perioperative safety in older adults. Excessive propofol exposure during induction may contribute to post-induction hypotension^8,23^ and may also increase the likelihood of EEG burst suppression, a marker of excessive anesthetic effect that has been associated with vulnerable brain states and adverse postoperative outcomes^24–26^. Although the *Ce*_50_ benchmarks used here should not be interpreted as prescriptive individual targets, they provide a physiologically grounded reference against which current practice can be evaluated. The magnitude of the observed exposure mismatch supports greater attention to tools that more directly reflect hypnotic effect, including neurophysiologic monitoring, rather than reliance on weight-based induction heuristics alone.

### Robustness across model and analytic choices

The overall pattern was consistent across multiple sensitivity analyses, suggesting that the identified age-related exposure mismatch is not dependent on a single modeling choice. Similar findings were obtained with the Schnider model despite its different treatment of age-related volumes and clearances, and the signal persisted across alternate analytic specifications, including analyses with less restrictive filtering and analyses including the masked *≥* 90-year-old cohort. In addition, adjustment for co-administered induction medications such as fentanyl, ketamine, midazolam, and etomidate did not materially change the central findings. Collectively, these analyses support the internal robustness of the observed age-related divergence between dose reduction and modeled exposure reduction.

### Limitations

Several limitations warrant consideration. First, this was a retrospective, model-based analysis and therefore depends on the accuracy of medication timestamps and the validity of the pharmacokinetic models used. Documentation error could affect individual estimates, although it is unlikely to explain the consistent population-level trends observed at this scale. Second, the *Ce*_50_ reference represents a population-average benchmark rather than an individualized target. It was used here as a standardized physiologic comparator rather than as a recommendation for bedside titration. Third, although some prior work has suggested possible model bias in elderly patients^27^, the qualitative findings were similar across different model structures, supporting the conclusion that the observed pattern is not solely a feature of one parameterization. Finally, this was a single-center study, and practice patterns may differ across institutions.

### Conclusions

In this cohort of more than 250,000 anesthetic inductions, age-related reductions in propofol dose were not matched by proportional reductions in modeled peak brain exposure. These findings suggest that weight-normalized induction dosing may inadequately reflect the pharmacology of aging and may underestimate the degree of exposure delivered to older adults. An exposure-based framework may provide a more informative way to evaluate and refine induction practice in the geriatric population.

## AUTHOR CONTRIBUTIONS

Brent Ershoff: Conceptualization, methodology, software, data curation, formal analysis, validation, visualization, writing–original draft, writing–review and editing.

## DATA AVAILABILITY

Code used for the pharmacokinetic simulations and statistical analyses is publicly available in a version-controlled GitHub repository at https://github.com/brentershoff/Propofol-. Due to institutional and privacy restrictions, the source perioperative electronic health record data cannot be shared publicly. A version corresponding to the final accepted manuscript will be archived upon acceptance.

## DECLARATION OF GENERATIVE AI AND AI-ASSISTED TECHNOLOGIES IN THE MANUSCRIPT PREPARATION PROCESS

During the preparation of this work, the author used ChatGPT (OpenAI) and Gemini (Google) in order to improve readability, language, and organization of the manuscript, and to assist with code drafting and code refinement. After using these tools/services, the author reviewed and edited the content as needed and takes full responsibility for the content of the published article.

## Data Availability

The data analyzed in this study were derived from a de-identified extract of the UCLA perioperative data warehouse. Although the dataset is de-identified, institutional policy and data-sharing agreements prohibit the public release of the raw underlying records.

https://github.com/brentershoff/Propofol-

## SUPPLEMENTARY MATERIAL

**Table S1:**
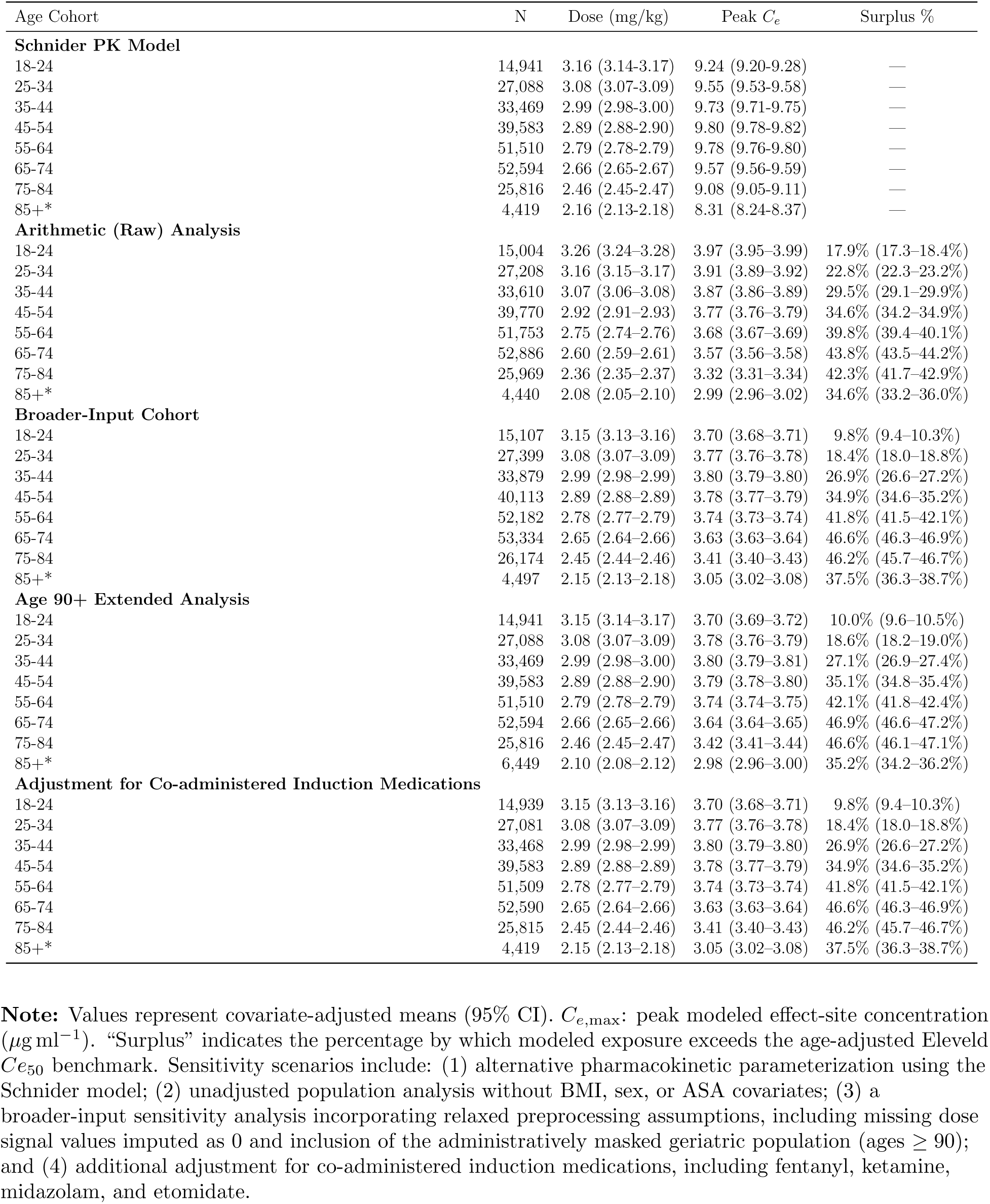
Robustness of age-associated titration offsets across alternative modeling and sensitivity-analysis specifications.

**Figure S1:**
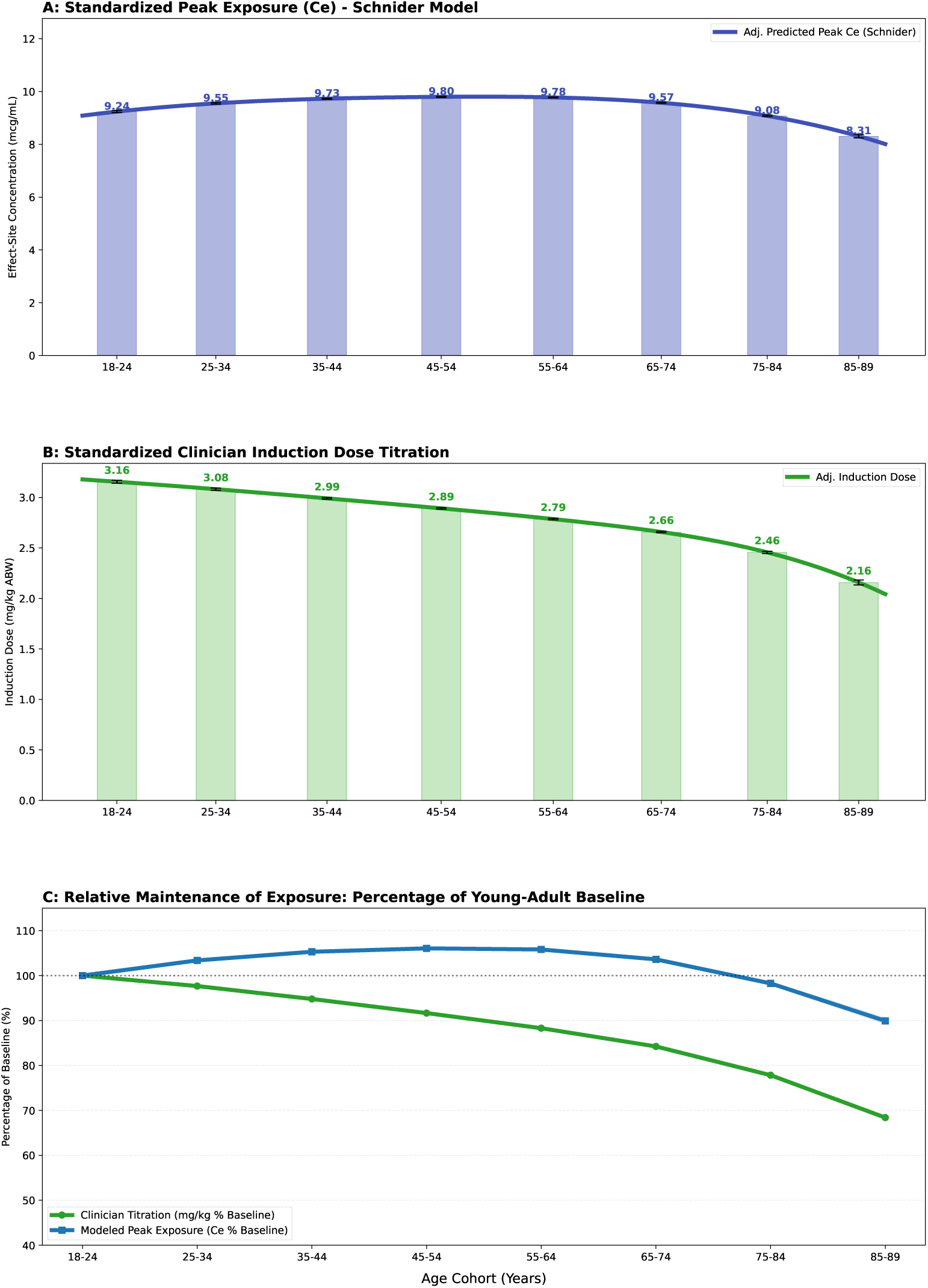
Sensitivity analysis using the Schnider pharmacokinetic model. Secondary analysis evaluating whether the age-associated exposure mismatch was sensitive to pharmacokinetic model specification. **(A) Peak effect-site exposure:** Covariate-adjusted predicted peak effect-site concentrations under the Schnider model across the adult lifespan. **(B) Relative divergence:** Normalized relative change in

**Figure S2:**
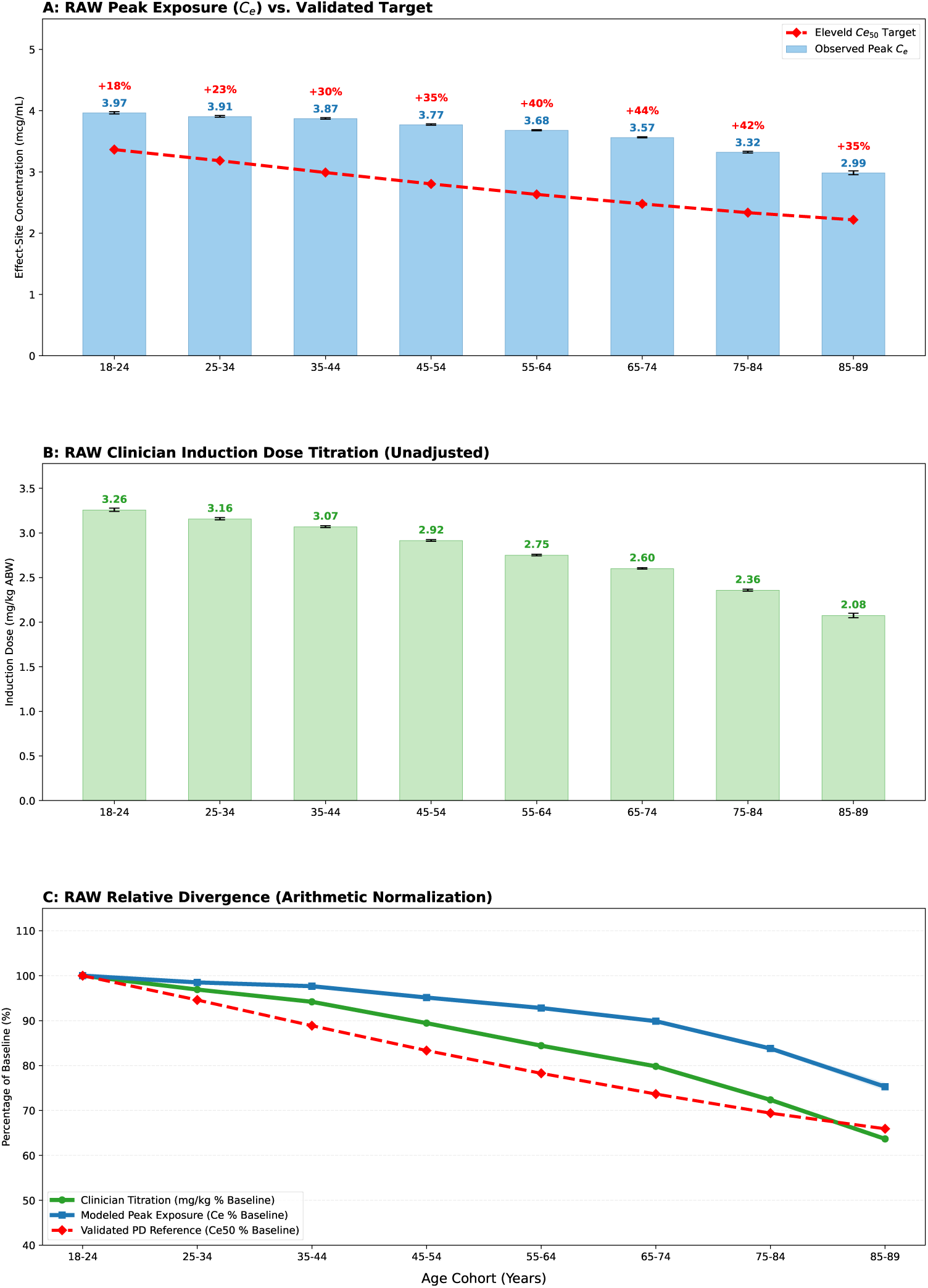
Unadjusted population-level analysis of propofol induction dynamics. Unadjusted arithmetic means derived directly from the EHR illustrate the age-associated divergence between clinical dosing, modeled peak exposure, and pharmacodynamic requirement before covariate standardization. **(A) Peak effect-site exposure versus requirement:** Observed peak modeled *Ce* (blue) shown alongside the age-adjusted Eleveld *Ce*_50_ benchmark (red). **(B) Clinician induction dose titration:** Observed weight-normalized induction doses (mg kg*^−^*^1^ ABW) across the adult lifespan. **(C) Relative divergence:** Normalized relative change in clinical dosing, modeled peak exposure, and pharmacodynamic requirement, indexed to the 18–24-year-old baseline. Error bars denote 95% confidence intervals.

**Figure S3:**
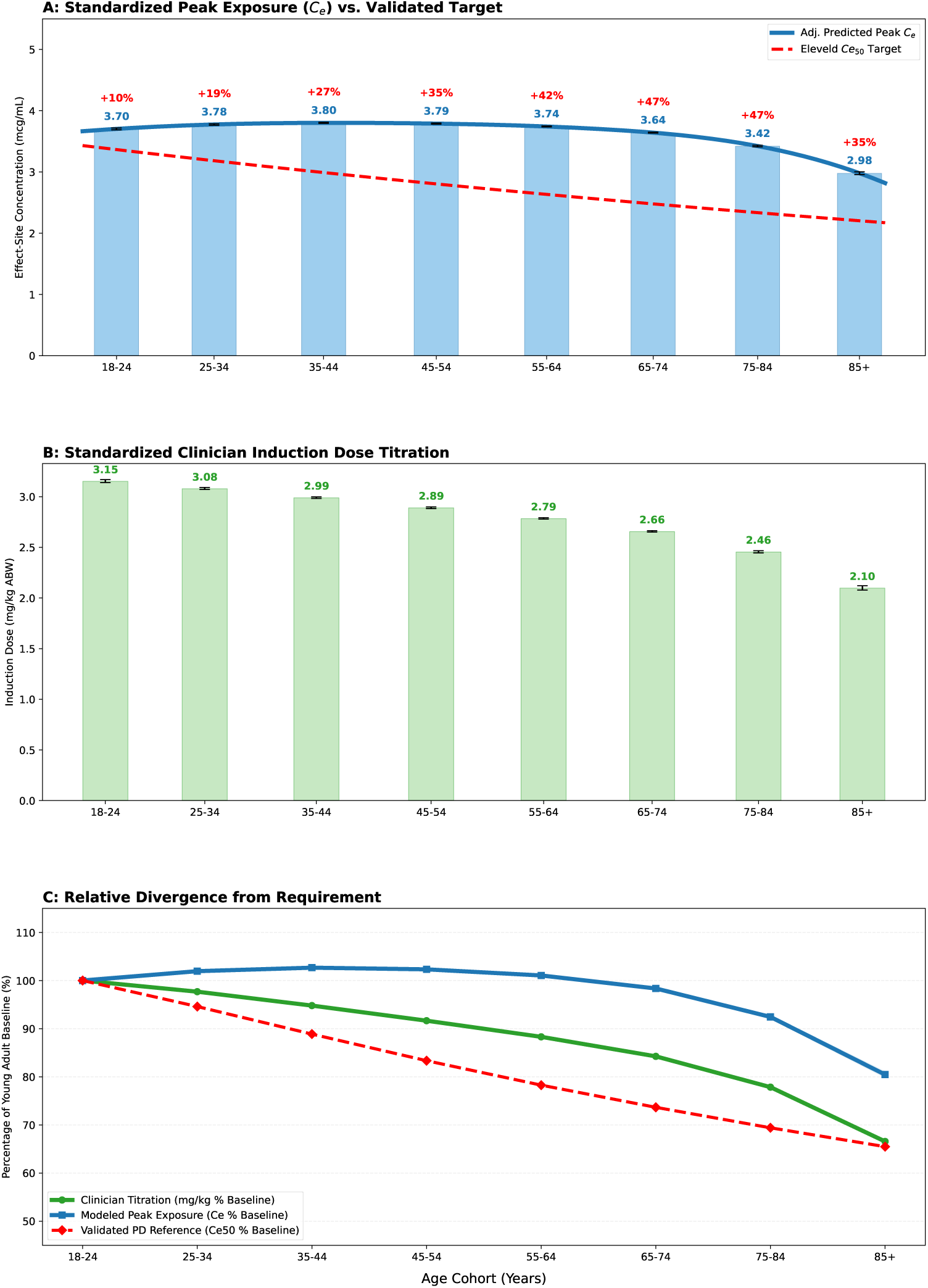
Sensitivity Analysis Including Patients Aged 90 and Older Standardized population-level dynamics including the advanced geriatric population. **(A) Standardized Peak Exposure (***C_e_*): Observed exposure (blue) versus Eleveld *Ce*_50_ requirement (red). **(B) Standardized Dose Titration:** Administered induction doses (mg kg*^−^*^1^) across the full lifespan. **(C) Standardized Relative Divergence:** Error bars denote 95% CI.

**Figure S4:**
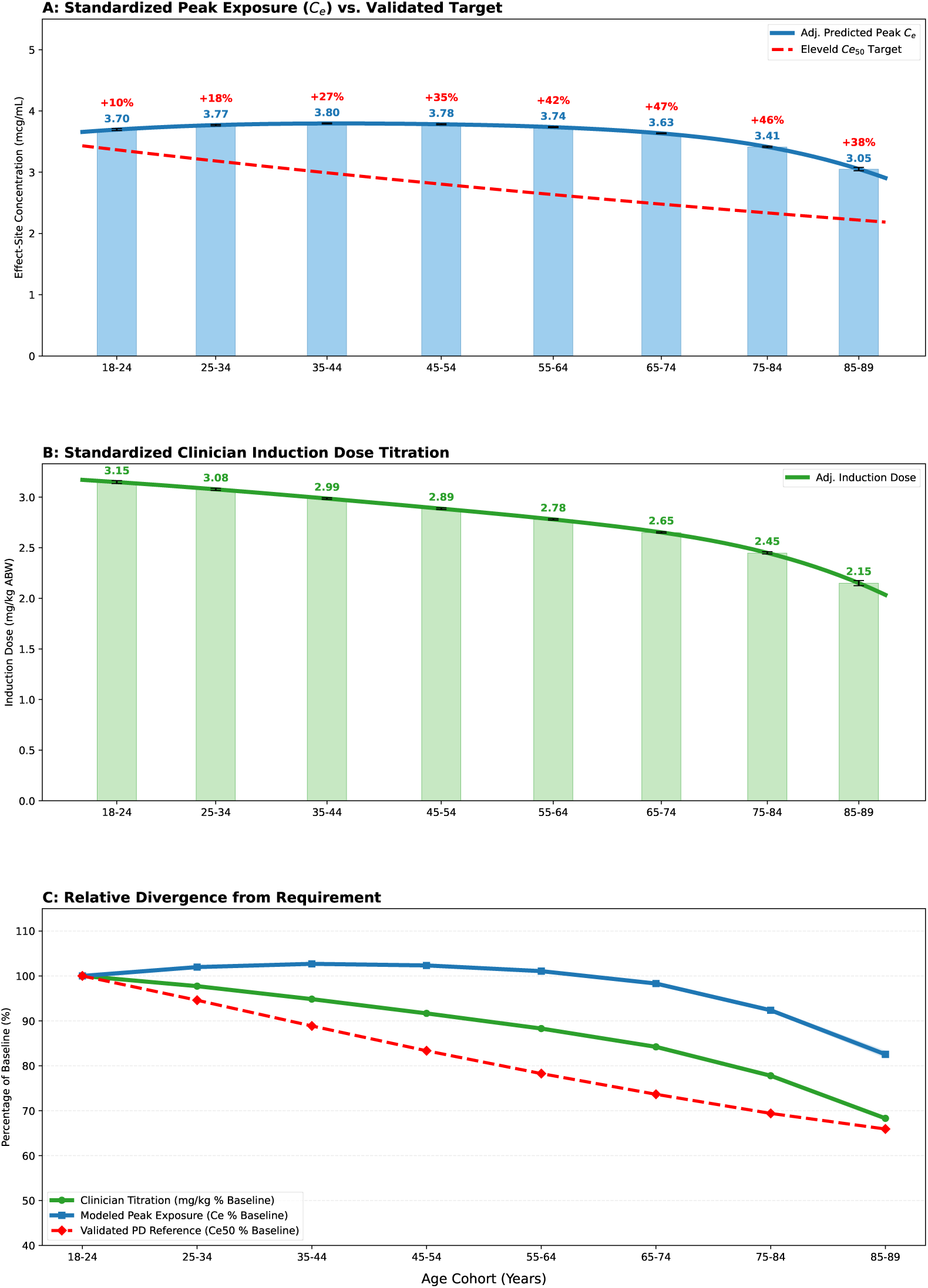
Broader-input sensitivity analysis with relaxed preprocessing assumptions. (A) Peak effect-site exposure versus requirement: Covariate-adjusted predicted peak effect-site concentrations (*C_e,_*_max_) shown alongside the age-adjusted Eleveld *Ce*_50_ benchmark. **(B) Clinician induction dose titration:** Covariate-adjusted predicted weight-normalized induction doses (mg kg*^−^*^1^ ABW) across the adult lifespan. **(C) Relative divergence:** Normalized relative change in clinical dosing, modeled peak exposure, and pharmacodynamic requirement, indexed to the 18–24-year-old baseline. This sensitivity analysis incorporated relaxed preprocessing assumptions, including missing dose signal values imputed as 0 and broader dose and physiologic inclusion thresholds. The persistence of the age-associated exposure mismatch indicates robustness to alternate preprocessing assumptions.

**Figure S5:**
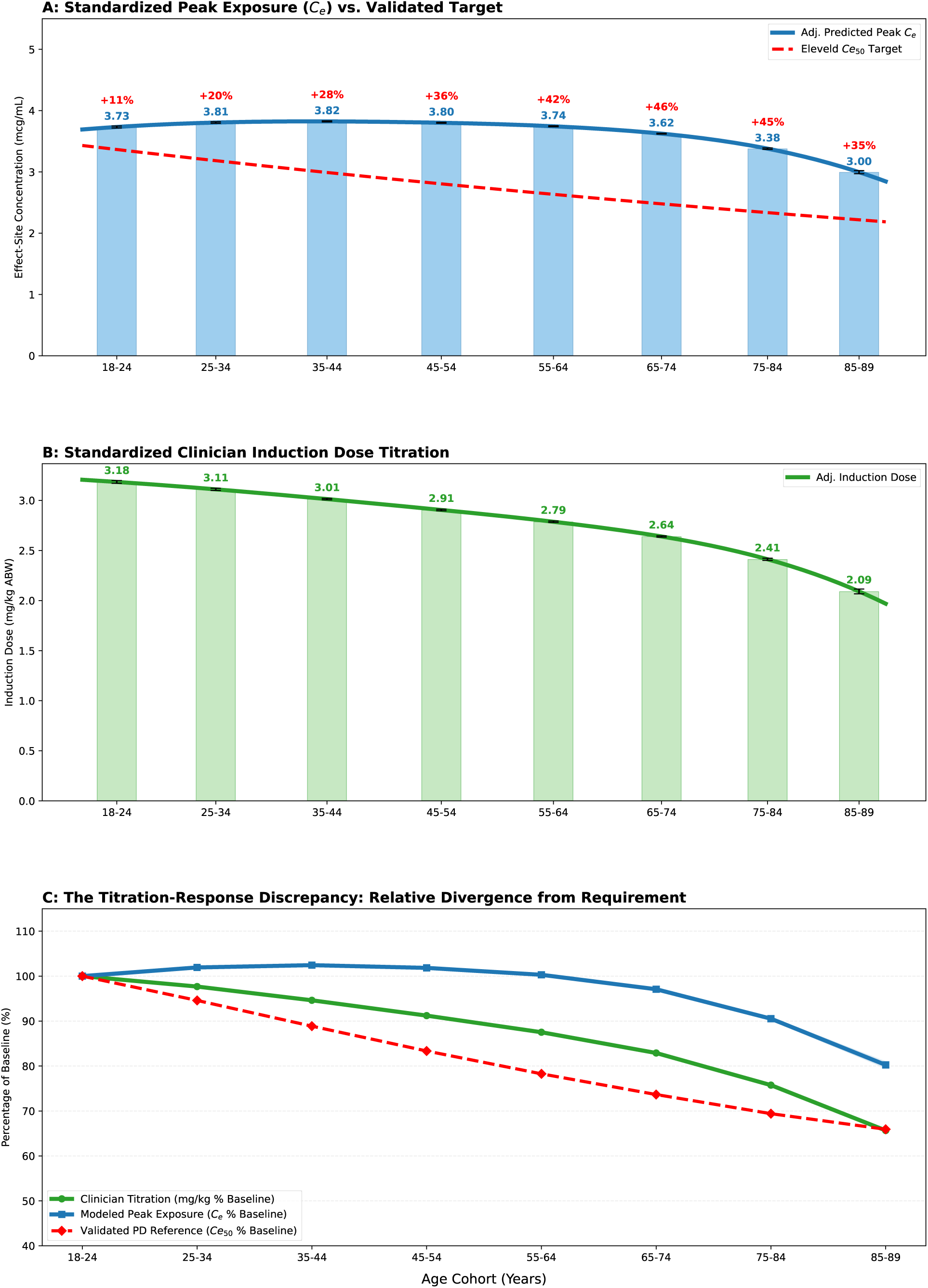
Adjunct-adjusted multidimensional analysis of propofol induction dynamics. **Figure S5: Adjunct-adjusted divergence of clinical dosing from modeled brain exposure and age adjusted pharmacodynamic requirement. (A) Exposure versus requirement:** Covariate-adjusted predicted peak effect-site concentrations (*C_e,_*_max_) shown alongside age-adjusted *Ce*_50_ benchmarks. **(B) Clinical dosing trajectory:** Covariate-adjusted predicted weight-normalized induction doses (mg kg*^−^*^1^ ABW) across the adult lifespan. **(C) Normalized relative divergence:** Relative change in clinical dosing (green), modeled exposure (blue), and pharmacodynamic requirement (red), indexed to the young-adult baseline (ages 18–24). **Note:** Curves represent adjusted predicted means from spline-based regression models, with BMI, ASA physical status, sex, and co-administered induction medications held at population means. Co-administered medications included fentanyl, ketamine, midazolam, and etomidate. Bars and error bars denote age-stratified adjusted predictions with 95% confidence intervals. In panel C, the blue confidence band denotes the 95% confidence interval for normalized modeled exposure. The dashed red line indicates the age-adjusted *Ce*_50_ benchmark from the propofol-only Eleveld model. Percentage labels indicate mean exposure surplus relative to the pharmacodynamic benchmark.

### SUPPLEMENTARY APPENDIX

#### Purpose of this appendix

This appendix provides additional technical and reproducibility details that extend, but do not replace, the Methods described in the main manuscript. The primary study design, cohort definition, pharmacokinetic reconstruction strategy, statistical framework, and benchmark definitions are reported in the main text. The sections below provide expanded implementation details, sensitivity-analysis specifications, validation notes, and code-availability information.

##### S1. Cohort construction and data extraction

Adult surgical cases receiving general anesthesia with propofol were identified from the UCLA perioperative data warehouse. The initial extraction yielded 258,741 cases. Sequential exclusions were then applied to remove records with implausible induction dosing, missing dose documentation, missing simulation variables, or physiologic outliers, as summarized in Figure 1 of the main manuscript.

In the primary analytic workflow, cases were sequentially excluded if they had missing dose signal values (SIG), missing critical simulation variables (AGE, WEIGHT KG, HEIGHT IN, SEX, ACTION TIME, UNITS, or BMI), invalid sex coding outside male/female categories, physiologic outliers defined by height outside 40–90 inches or BMI outside 8–100, or total propofol dose outside 20–700 mg. When height and weight were available, missing BMI values were first recalculated before attrition. Cases with age *≥* 90 years were excluded from the primary analysis because the source system administratively records all such patients as age 90 for de-identification, preventing use of exact chronological age in age-based pharmacokinetic and pharmacodynamic modeling. A secondary sensitivity analysis re-incorporated this administratively masked cohort to evaluate whether exclusion of these cases materially affected the primary findings.

Propofol medication records were extracted using Structured Query Language (SQL) from the institutional perioperative data warehouse. The cohort was restricted to cases in which the documented induction event preceded the documented intubation timestamp. For each included case, the induction timestamp served as the reference point for medication extraction. To capture the dynamic induction phase, all propofol administrations occurring within 10 minutes after the induction event were retrieved. Extracted records included both discrete bolus doses (mg) and continuous infusion events, including infusion start times, stop times, and rate changes (*µ*g kg^−1^ min^−1^). Each medication record retained its source timestamp and dosing units for downstream pharmacokinetic reconstruction.

To contextualize modeled effect-site exposure against conventional clinical dosing metrics, total induction dose was normalized using adjusted body weight (ABW). Ideal body weight (IBW) was calculated using the Devine formula. For patients whose total body weight (TBW) exceeded 120% of IBW, ABW was calculated as

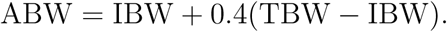

For patients whose TBW did not exceed 120% of IBW, TBW was used as the dosing weight. Total propofol administered within the induction extraction window was then divided by ABW to generate the weight-normalized induction dose (mg kg^−1^).

##### S2. Pharmacokinetic reconstruction and exposure benchmarking

Effect-site concentration time series were reconstructed by simulating validated propofol PK/PD models forward in time using the observed medication event log for each case as input. Simulations were implemented using the open-source PyTCI library.^15^ The Eleveld model served as the primary pharmacokinetic framework because it incorporates age, sex, height, and weight across a broad range of patient characteristics represented in the study cohort.^13^

For each case, all medication events were aligned relative to the first documented propofol administration, which was defined as *t* = 0. Subsequent event times were represented in integer seconds relative to this first dose. Simulations were then run at 1-second resolution for a fixed 900-second horizon beginning at *t* = 0. Between medication events, the PK/PD model state was advanced in 1-second increments. Bolus doses recorded in milligrams were applied instantaneously at the documented administration time. Infusion events recorded in (*µ*g kg^−1^ min^−1^)in were converted to (mg s^−1^) and applied continuously until modified by a subsequent documented rate change or infusion stop event. At each simulation time point, the evolving plasma and effect-site concentrations were updated according to the selected model equations. The primary exposure metric was the peak modeled effect-site concentration within the 900-second simulation window,

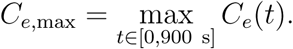

To provide a standardized physiologic benchmark for age-related hypnotic requirement, modeled peak effect-site concentrations were compared with the age-adjusted *Ce*_50_ relationship derived from the propofol-only Eleveld model:

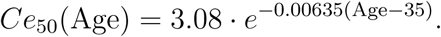

In the propofol-only configuration, this relationship corresponds to the modeled effect-site concentration associated with a Bispectral Index (BIS) value of 47 and was used in the present study as a population-average reference rather than an individualized bedside target. The purpose of this benchmark was to provide a consistent age-dependent slope against which achieved modeled exposure could be compared across the adult lifespan.

Divergence between observed clinical dosing patterns and age-adjusted pharmacodynamic requirement was characterized in three ways: (1) relative normalization of *C_e,_*_max_, weight-normalized dose, and *Ce*_50_ to the 18–24-year-old cohort; (2) estimation of the probability that *C_e,_*_max_ *> Ce*_50_(Age); and (3) estimation of the probability that *C_e,_*_max_ exceeded the model-predicted *Ce*_50_ for a 21-year-old patient (3.37 *µ*g ml^−1^), used as a young-adult benchmark.

##### S2.1 Simulation algorithm for Eleveld-based effect-site reconstruction

To improve transparency of the event-driven pharmacokinetic reconstruction, the primary Eleveld-based simulation workflow is summarized in Algorithm 1. This pseudocode is intended to illustrate the conceptual implementation used to convert retrospective bolus and infusion records into modeled effect-site concentration trajectories and patient-level peak exposure estimates.

**Algorithm 1.**
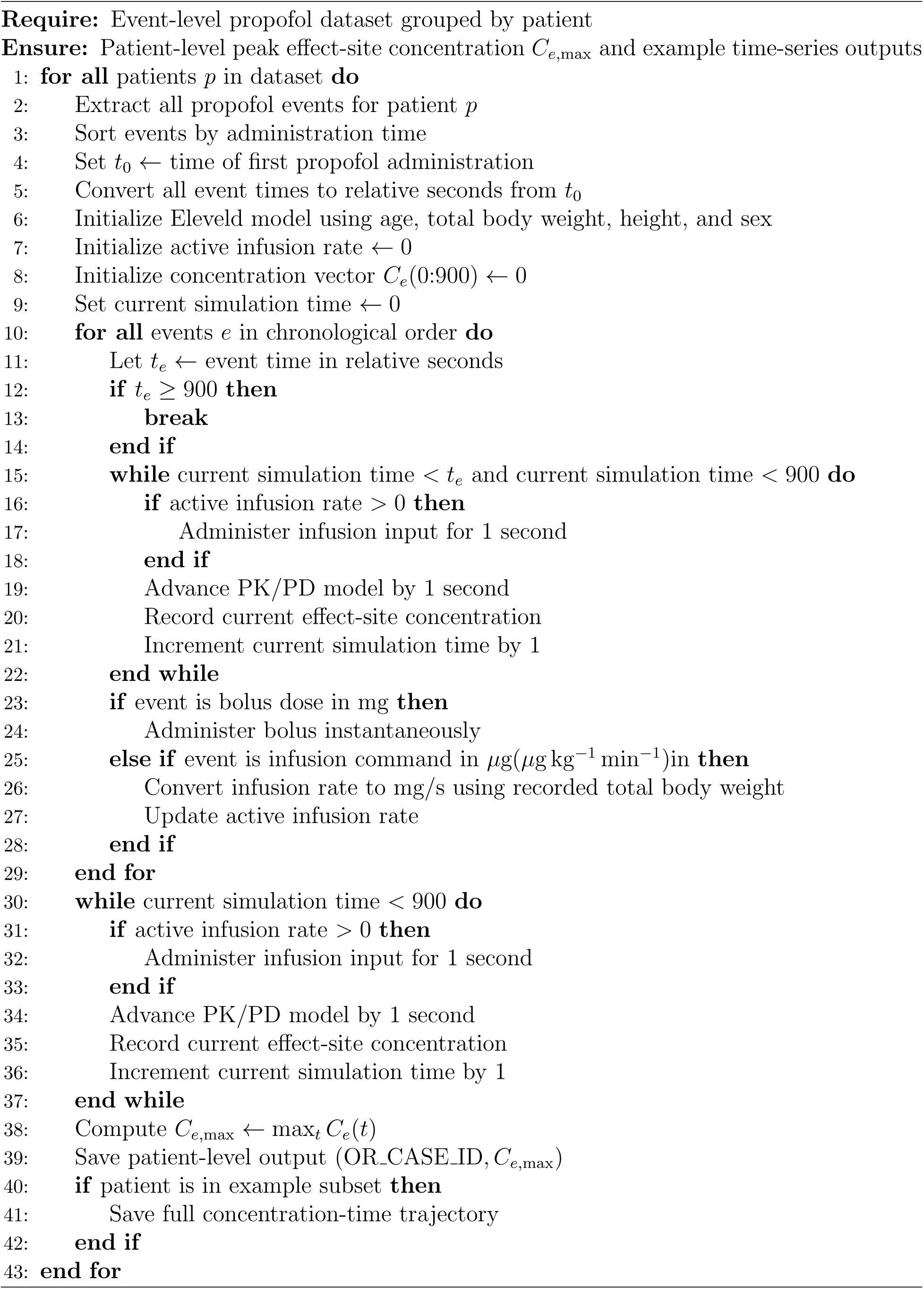
Eleveld-based retrospective effect-site concentration reconstruction.

Algorithm 1 summarizes the primary reconstruction logic used in the manuscript. The Schnider sensitivity analysis used the same general event-driven framework, but substituted the Schnider propofol model and its corresponding model-specific parameterization. Full executable implementations of both the primary and sensitivity-model workflows are provided in the public code repository and archived release associated with this manuscript.

##### S3. Statistical analyses and sensitivity analyses

All statistical analyses were performed in Python. Data wrangling and numerical computation used pandas and NumPy, and regression modeling used statsmodels with spline design matrices generated using patsy.^17–21^

The association between age and peak modeled effect-site concentration (*C_e,_*_max_) was estimated using ordinary least squares regression with age modeled as a B-spline with five degrees of freedom. An analogous approach was used to model age-associated changes in weight-normalized induction dose (mg kg^−1^ ABW). Overexposure outcomes were modeled using logistic regression with age represented by B-spline terms and adjustment for sex, BMI, and ASA physical status. Covariate-adjusted predictions were obtained using marginal prediction for a standardized patient profile in which BMI, ASA physical status, and sex were fixed at cohort-level means. For figure presentation, model-predicted values and 95% confidence intervals were evaluated both continuously across age and at the mean age of each prespecified age stratum (18–24, 25–34, 35–44, 45–54, 55–64, 65–74, 75–84, and 85–89 years).

Several sensitivity analyses were performed to evaluate the robustness of the primary findings. These included repetition of the full pharmacokinetic reconstruction workflow using the Schnider propofol model,^16^ re-estimation of primary age trajectories without adjustment for BMI, sex, or ASA physical status, and inclusion of the administratively masked age-*≥* 90 cohort. A broader-input sensitivity analysis was also performed in which missing dose signal values (SIG) were imputed as 0 rather than excluded, missing BMI values were recalculated from height and weight when possible, and cases were retained provided they had non-missing core simulation variables, valid sex coding, height between 40–90 inches, BMI between 8–200, and total propofol dose between 0–2000 mg. In addition, regression models were re-estimated with adjustment for fentanyl, ketamine, midazolam, and etomidate administered during the induction window. To limit the influence of implausible or extreme co-medication values, this sensitivity analysis excluded cases with etomidate *>* 60 mg, fentanyl *>* 1000 *µ*g, or ketamine *>* 300 mg, removing 16 patients before modeling. Age-stratified estimates and confidence intervals for all sensitivity analyses are provided in the supplementary tables and figures.

##### S4. Validation, code availability, and repository structure

To assess the face validity of the automated pharmacokinetic reconstruction pipeline, representative induction scenarios were independently recreated in TivaTrainer.^22^ These scenarios included both isolated bolus administrations and combined bolus-plus-infusion sequences. Peak effect-site concentrations and full concentration-time trajectories generated by the study simulation engine were then compared with the corresponding TivaTrainer outputs. Near-identical agreement between the reconstructed trajectories and external TivaTrainer benchmarks supported the fidelity of the event parsing, time alignment, unit conversion, and model implementation used in the retrospective simulation framework.

#### Repository organization

The public repository contains the code used for cohort extraction, pharmacokinetic simulation, statistical modeling, and generation of the main and supplementary figures and tables. The repository README describes the execution order of notebooks, software dependencies, and the correspondence between analytic files and manuscript outputs.

**Table S2:**
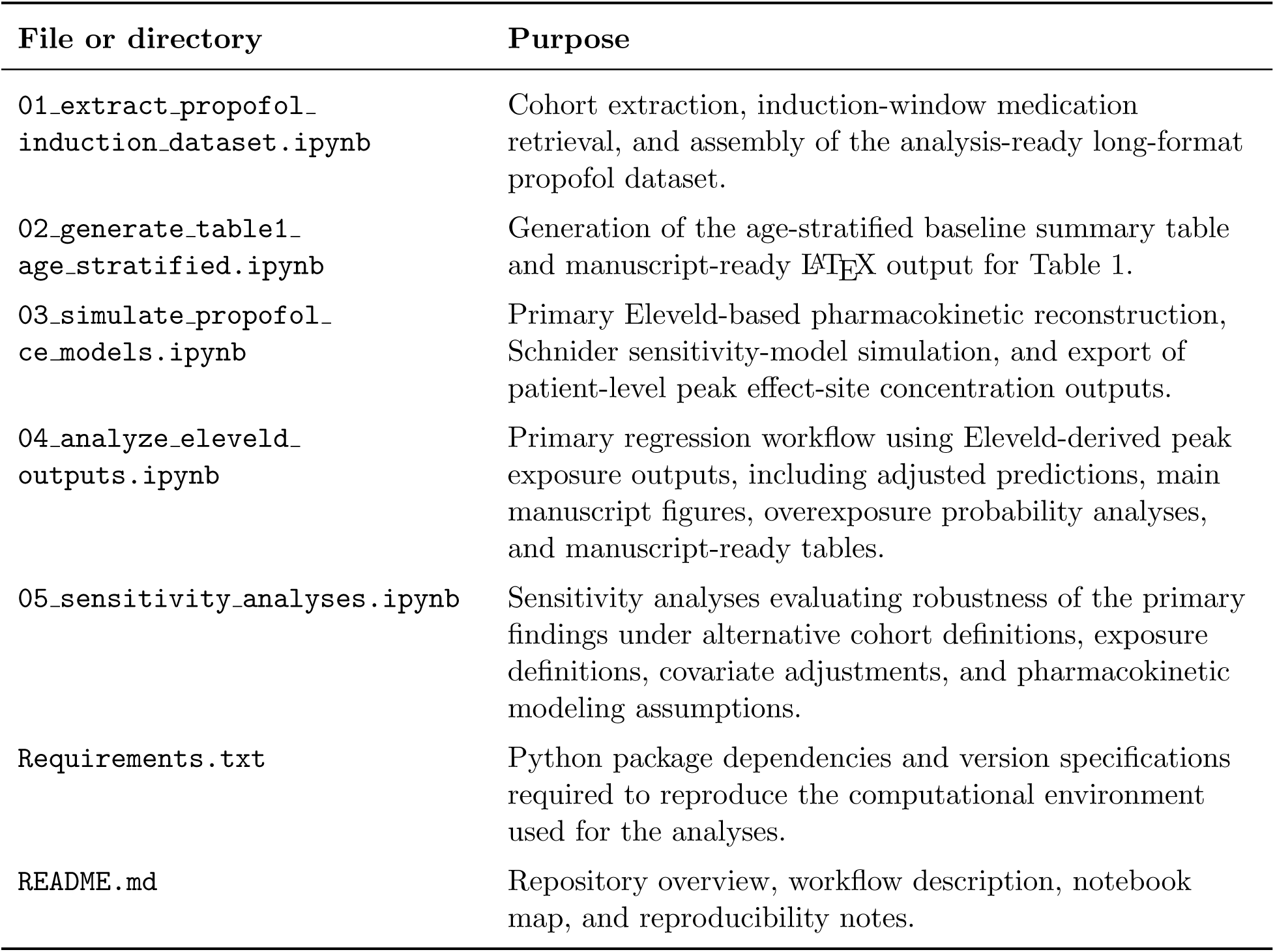
Repository organization and correspondence to manuscript outputs.

| File or directory | Purpose |
| --- | --- |
| 01_extract_propofol_induction_dataset.ipynb | Cohort extraction, induction-window medication retrieval, and assembly of the analysis-ready long-format propofol dataset. |
| 02_generate_table1_age_stratified.ipynb | Generation of the age-stratified baseline summary table and manuscript-ready L<sup>A</sup>T<sub>E</sub>X output for Table 1. |
| 03_simulate_propofol_ce_models.ipynb | Primary Eleveld-based pharmacokinetic reconstruction, Schnider sensitivity-model simulation, and export of patient-level peak effect-site concentration outputs. |
| 04_analyze_eleveld_outputs.ipynb | Primary regression workflow using Eleveld-derived peak exposure outputs, including adjusted predictions, main manuscript figures, overexposure probability analyses, and manuscript-ready tables. |
| 05_sensitivity_analyses.ipynb | Sensitivity analyses evaluating robustness of the primary findings under alternative cohort definitions, exposure definitions, covariate adjustments, and pharmacokinetic modeling assumptions. |
| Requirements.txt | Python package dependencies and version specifications required to reproduce the computational environment used for the analyses. |
| README.md | Repository overview, workflow description, notebook map, and reproducibility notes. |

## References

1. Prince, M. J., Wu, F., Guo, Y., Gutierrez Robledo, L. M., O’Donnell, M., Sullivan, R. & Yusuf, S. The burden of disease in older people and implications for health policy and practice. The Lancet 385, 549–562 (Feb. 2015).

2. Deiner, S., Westlake, B. & Dutton, R. P. Patterns of Surgical Care and Complications in Elderly Adults. Journal of the American Geriatrics Society 62, 829–835 (May 2014).

3. Becher, R. D., Vander Wyk, B., Leo-Summers, L., Desai, M. M. & Gill, T. M. The Incidence and Cumulative Risk of Major Surgery in Older Persons in the United States. Annals of Surgery 277, 87–92 (Jan. 2023).

4. Yang, H., Deng, H.-M., Chen, H.-Y., Tang, S.-H., Deng, F., Lu, Y.-G. & Song, J.-C. The Impact of Age on Propofol Requirement for Inducing Loss of Consciousness in Elderly Surgical Patients. Frontiers in Pharmacology 13, 739552 (Mar. 28, 2022).

5. Schnider, T. W., Minto, C. F., Shafer, S. L., Gambus, P. L., Andresen, C., Goodale, D. B. & Youngs, E. J. The Influence of Age on Propofol Pharmacodynamics. Anesthesiology 90, 1502–1516. (June 1, 1999).

6. Propofol Injection, Emulsion [package insert]. U.S. Food and Drug Administration https://fda.report/DailyMed/fdb77e13-f5a7-4c4a-9f8e-338b702239c8/. Accessed February 20, 2026 (U.S. Food and Drug Administration).

7. Schonberger, R. B., Bardia, A., Dai, F., Michel, G., Yanez, D., Curtis, J. P., Vaughn, M. T., Burg, M. M., Mathis, M., Kheterpal, S., Akhtar, S. & Shah, N. Variation in Propofol Induction Doses Administered to Surgical Patients over Age 65. Journal of the American Geriatrics Society 69, 2195–2209 (Aug. 2021).

8. Phillips, A. T., Deiner, S., Lin, H. M., Andreopoulos, E., Silverstein, J. & Levin, M. A. Propofol Use in the Elderly Population: Prevalence of Overdose and Association With 30-Day Mortality. Clinical therapeutics 37, 2676–2685 (Dec. 1, 2015).

9. Akhtar, S., Liu, J., Heng, J., Dai, F., Schonberger, R. B. & Burg, M. M. Does intravenous induction dosing among patients undergoing gastrointestinal surgical procedures follow current recommendations: a study of contemporary practice. Journal of Clinical Anesthesia 33, 208–215 (Sept. 2016).

10. Chen, E. Y., Michel, G., Zhou, B., Dai, F., Akhtar, S. & Schonberger, R. B. An Analysis of Anesthesia Induction Dosing in Female Older Adults. Drugs & Aging 37, 435–446 (June 2020).

11. Al-Rifai, Z. & Mulvey, D. Principles of total intravenous anaesthesia: practical aspects of using total intravenous anaesthesia. BJA Education 16, 276–280 (Aug. 2016).

12. Vandemoortele, O., Hannivoort, L. N., Vanhoorebeeck, F., Struys, M. M. R. F. & Vereecke, H. E. M. General Purpose Pharmacokinetic-Pharmacodynamic Models for Target-Controlled Infusion of Anaesthetic Drugs: A Narrative Review. Journal of Clinical Medicine 11, 2487 (Apr. 28, 2022).

13. Eleveld, D., Colin, P., Absalom, A. & Struys, M. Pharmacokinetic–pharmacodynamic model for propofol for broad application in anaesthesia and sedation. British Journal of Anaesthesia 120, 942–959 (May 2018).

14. Hofer, I. S., Gabel, E., Pfeffer, M., Mahbouba, M. & Mahajan, A. A Systematic Approach to Creation of a Perioperative Data Warehouse. Anesthesia & Analgesia 122, 1880–1884 (June 2016).

15. Mathiszig-Lee, J. PyTCI: Open-source Target-Controlled Infusion simulation library. https://github.com/JMathiszig-Lee/PyTCI. 2022.

16. Schnider, T. W., Minto, C. F., Gambus, P. L., Andresen, C., Goodale, D. B., Shafer, S. L. & Youngs, E. J. The Influence of Method of Administration and Covariates on the Pharmacokinetics of Propofol in Adult Volunteers. Anesthesiology 88, 1170–1182 (May 1, 1998).

17. Van Rossum, G. & Drake, F. *Python Programming Language* version 3.12.12. 2009.

18. McKinney, W. *Data Structures for Statistical Computing in Python* in. Python in Science Conference (Austin, Texas, 2010), 56–61.

19. Harris, C. R., Millman, K. J., Van Der Walt, S. J., Gommers, R., Virtanen, P., Cournapeau, D., Wieser, E., Taylor, J., Berg, S., Smith, N. J., Kern, R., Picus, M., Hoyer, S., Van Kerkwijk, M. H., Brett, M., Haldane, A., Del Río, J. F., Wiebe, M., Peterson, P., Gérard-Marchant, P., Sheppard, K., Reddy, T., Weckesser, W., Abbasi, H., Gohlke, C. & Oliphant, T. E. Array programming with NumPy. Nature 585, 357–362 (Sept. 17, 2020).

20. Seabold, S. & Perktold, J. *Statsmodels: Econometric and Statistical Modeling with Python* in. Python in Science Conference (Austin, Texas, 2010), 92–96.

21. Smith, N. J., Broessli, Skipper Seabold & Davidson-Pilon, C. patsy v0.2.1 version v0.2.1. Aug. 26, 2014.

22. Engbers, F. *TIVAtrainer* version 3.1.1. 2026.

23. Dundee, J. W., Robinson, F. P., McCOLLUM, J. S. C. & Patterson, C. C. Sensitivity to propofol in the elderly. Anaesthesia 41, 482–485 (May 1986).

24. Obert, D. P., Sepúlveda, P. O., Adriazola, V., Zurita, F., Brouse, J., Schneider, G. & Kreuzer, M. Overcoming age: Slow anesthesia induction may prevent geriatric patients from developing burst suppression and help developing intraoperative EEG signatures of a younger brain. Journal of Clinical Anesthesia 99, 111672 (Dec. 1, 2024).

25. Purdon, P., Pavone, K., Akeju, O., Smith, A., Sampson, A., Lee, J., Zhou, D., Solt, K. & Brown, E. The Ageing Brain: Age-dependent changes in the electroencephalogram during propofol and sevoflurane general anaesthesia. British Journal of Anaesthesia 115, i46–i57 (July 2015).

26. Fritz, B. A., King, C. R., Ben Abdallah, A., Lin, N., Mickle, A. M., Budelier, T. P., Oberhaus, J., Park, D., Maybrier, H. R., Wildes, T. S., Avidan, M. S. & for the ENGAGES Research Group*. Preoperative Cognitive Abnormality, Intraoperative Electroencephalogram Suppression, and Postoperative Delirium: A Mediation Analysis. Anesthesiology 132, 1458–1468 (June 2020).

27. Vellinga, R., Hannivoort, L. N., Introna, M., Touw, D. J., Absalom, A. R., Eleveld, D. J. & Struys, M. M. F. Prospective clinical validation of the Eleveld propofol pharmacokinetic-pharmacodynamic model in general anaesthesia. British Journal of Anaesthesia 126, 386–394 (Feb. 2021).

